# Molecular epidemiology of SARS-CoV-2 in Greece reveals low rates of onward virus transmission after lifting of travel restrictions based on risk assessment during summer 2020

**DOI:** 10.1101/2021.01.31.21250868

**Authors:** Evangelia Georgia Kostaki, Georgios A. Pavlopoulos, Kleio-Maria Verrou, Giannis Ampatziadis-Michailidis, Vaggelis Harokopos, Pantelis Hatzis, Panagiotis Moulos, Nikolaos Siafakas, Spyridon Pournaras, Christos Hadjichristodoulou, Fani Chatzopoulou, Dimitrios Chatzidimitriou, Periklis Panagopoulos, Panagiota Lourida, Aikaterini Argyraki, Theodoros Lytras, Spyros Sapounas, Gerasimos Gerolymatos, Georgios Panagiotakopoulos, Panagiotis Prezerakos, Sotirios Tsiodras, Vana Sypsa, Angelos Hatzakis, Cleo Anastassopoulou, Nikolaos Spanakis, Athanasios Tsakris, Meletios Athanasios Dimopoulos, Anastasia Kotanidou, Petros Sfikakis, Georgios Kollias, Gkikas Magiorkinis, Dimitrios Paraskevis

**Affiliations:** Department of Hygiene, Epidemiology and Medical Statistics, Medical School, National and Kapodistrian University of Athens, Athens, Greece; Center of New Biotechnologies & Precision Medicine, Medical School, National and Kapodistrian University of Athens, Athens, Greece; Institute for Fundamental Biomedical Research, Biomedical Sciences Research Center ‘Alexander Fleming’, Vari, Greece; Laboratory of Clinical Microbioloy, ATTIKON University Hospital, Medical School, National and Kapodistrian University of Athens, Athens, Greece; Laboratory of Hygiene and Epidemiology, Faculty of Medicine, Larisa, Greece; Labnet, Laboratories, Thessaloniki, Greece; Department of Microbiology, Medical School, Aristotle University of Thessaloniki, Thessaloniki, Greece; 2nd Department of Internal Medicine, General Hospital of Alexandroupoli, Democritus University of Thrace, Alexandroupoli, Greece; Infectious Diseases Clinic A, Thoracic Diseases General Hospital Sotiria, Athens, Greece; School of Medicine, European University Cyprus, Nicosia, Cyprus; National Public Health Organization, Athens, Greece; Department of Nursing, University of Peloponnese, Tripoli, Greece; 4th Department of Internal Medicine, Attikon University Hospital, Medical School, National and Kapodistrian University of Athens, Athens, Greece; Department of Microbiology, Medical School, National and Kapodistrian University of Athens, Athens, Greece; Department of Therapeutics, Medical School, National and Kapodistrian University of Athens, Athens, Greece; 1st Intensive Care Unit, General Hospital Evangelismos, National and Kapodistrian University of Athens, Athens, Greece; 1st Department of Prop. Internal Medicine, Medical School, National and Kapodistrian University of Athens, Athens, Greece; Institute for Bioinnovation, Biomedical Sciences Research Center ‘Alexander Fleming’, Vari, Greece; Department of Physiology, Medical School, National and Kapodistrian University of Athens, Athens, Greece

## Abstract

Molecular epidemiology has provided an additive value to traditional public health tools by identifying SARS-CoV-2 clusters, or providing evidence that clusters based on virus sequences and contact tracing are highly concordant. Our aim was to infer the levels of virus importation and to estimate the impact of public health measures related to travel restrictions to local transmission in Greece. Our phylogenetic and phylogeographic analyses included 389 SARS-CoV-2 sequences collected during the first 7 months of the pandemic in Greece and a random collection in 5 replicates of 3,000 sequences sampled globally, as well as the best hits to our dataset identified by BLAST. Phylogenetic analyses revealed the presence of 70 genetically distinct viruses identified as independent introductions into Greece. The proportion of imported strains was 41%, 11.5%, and 8.8% during the three periods of sampling, namely, March (no travel restrictions), April to June (strict travel restrictions), and July to September (lifting of travel restrictions based on a thorough risk assessment), respectively. These findings reveal low levels of onward transmission from imported cases during summer and underscore the importance of targeted public health measures that can increase the safety of international travel during a pandemic.

## Introduction

In December 2019, a new respiratory disease was described in Wuhan, China, which was found to be caused by a novel corona virus named Severe Acute Respiratory Syndrome coronavirus 2 (SARS-CoV-2) [1]. The new virus spread globally and caused a large pandemic associated with increased morbidity and mortality rates [2]. In the absence of an effective vaccine in the first year of the pandemic, non-pharmaceutical interventions (NPIs) such as social distancing, use of masks in the community, travel restrictions, school, and non-essential shops closures were implemented to control community transmission [3]. The health, social and economic consequences of the pandemic are continuous thus a better understanding of the characteristics of SARS-CoV-2 transmission is needed to minimize its consequences.

Molecular epidemiology analyses of SARS-CoV-2 full-genome sequences have been extensively performed to classify viral diversity into groups or lineages/sublineages [4], to provide continuous monitoring of the virus dispersal patterns and to obtain insights into critical epidemiological or public health issues related to the geographic origin and dating of viral transmission [5-7]. For example, phylogenetic studies revealed that the origin of transmission during the first pandemic wave in Italy and Seattle, Washington, were from different sources in Asia [8]. Phylogenetic analysis of virus samples revealed SARS-CoV-2 clusters and tourism associated virus dispersal of the first wave in Austria [9]. In the UK where virus genetic diversity has been systematically surveyed [5], a detailed description of the characteristics of transmission by means of the number and size of local clusters has been performed, as well as quantification of the spatiotemporal characteristics of viral diversity [10].The origin and dynamics of virus importation pattern during the first wave in the UK were also mapped [10]. Importantly a study from Iceland reported high concordance between the contacts identified by contact tracing and molecular data, suggesting that the latter can be used to control community viral transmission [11]. Recently, genomic surveillance has been of interest due to reports that certain new lineages found to rapidly spread across the UK, South Africa, Brazil and other areas in the recent months (i.e. B.1.1.7, B.1.351 and P.1) may confer different biological characteristics to the virus [12-14].

In Greece, the first pandemic wave was mild due to the early implementation of public health measures and the high compliance of the population to the imposed lock downs [15, 16]. However, by the end of October 2020 the country experienced rapid increases in the number of SARS-CoV-2 cases in the metropolitan area of Thessaloniki and other areas of Northern Greece. In the meantime, between the lifting of the first measures in May and this second wave, the number of cases remained relatively low even after travel restrictions were lifted at the beginning of July 2020. To date, several issues remain unanswered, such as how SARS-CoV-2 was introduced in the country, what is the pattern of virus dispersal and, importantly, what the impact of lifting of travel restrictions was on SARS-CoV-2 transmission

By applying molecular epidemiology methods, we aimed to quantify the levels of virus importation in comparison with surveillance data during these different time periods, to investigate the patterns of SARS-CoV-2 dispersal and to estimate the impact of public health measures related to travel restrictions to local transmission in Greece.

## Materials and methods

### Analysed SARS-CoV-2 samples

The SARS-CoV-2 samples analyzed in the context of the current study were collected from February 29 to September 19, 2020 in the Attica, Larisa and Thrace regions, from two SARS-CoV-2 reference centers. Specifically, all SARS-CoV-2 positive samples available with RT-PCR threshold detection cycles (Ct) ≤ 30 until the end of August 2020 from Attikon University Hospital (first Reference Center) were included in our analysis. Similarly, samples fulfilling the previous criterion available in September 2020 at the Department of Hygiene, Epidemiology and Medical Statistics of the School of Medicine at National and Kapodistrian University of Athens (second Reference Center) were included in our analysis. SARS-CoV-2 samples obtained at border control areas from travelers arriving in Greece were excluded from the analysis.

The study was approved by the Ethics and Bioethics Committee of the Medical School, of the National and Kapodistrian University of Athens (protocol #300/25-05-2020).

The SARS-CoV-2 samples selected were part of the routine diagnostic procedures performed in Greece at the two reference centers in Attica or elsewhere in Greece. SARS-CoV-2 testing in Greece is prioritized for symptomatic individuals, high-risk contacts of SARS-CoV-2 positive cases, or populations at high risk for SARS-CoV-2 infection (i.e., health care workers, residential nursing homes for elderly or disabled people). Samples collected in September included those performed by the National Public Health Organization (https://eody.gov.gr/en/) as part of volunteer testing of the population in Attica.

Our sampling comprised three time periods: between February 29 and March 31; between April 1 and June 30; and between July 1 and September 29, 2020. The three time periods were defined according to the status of travel restrictions implemented in Greece during the SARS-CoV-2 pandemic. Specifically, a travel ban and quarantine measures for all travellers were implemented on March 14 and 16, respectively, and, given the length of the SARS-CoV-2 incubation period, the first phase was extended until the end of March, 2020. The second period corresponded to when the international travel restrictions were in place, including a quarantine period for arriving subjects, and the third corresponded to the period after the lifting of travel restrictions based on a thorough risk assessment for all international travellers.

### Next-generation sequencing (NGS)

RNA samples were processed using the CleanPlex® SARS-CoV-2 Panel, (Paragon Genomics), according to manufacturer’s instructions. Samples were quantitated (Qubit™ RNA HS Assay Kit, Thermofisher), and 50-100 ng of total RNA was used for library preparation, with a final PCR amplification of 24-26 cycles. The resulting libraries were analyzed on a Bioanalyzer System (High Sensitivity DNA Kit, Agilent), quantified (Qubit dsDNA HS Assay Kit, Thermofisher), and multiplexed; they were sequenced on a NextSeq 550 System (Illumina), using the Mid Output Kit v2.5 (300 cycles), in paired end mode.

The quality of FASTQ files was assessed using FastQC (version 0.11.9) [17]. Adapter and poor-quality base trimming were performed with TrimGalore (version 0.6.6) [18], which also deploys Cutadapt (version 2.8) [19], keeping reads with length of at least 50 bp. Subsequently, the remaining paired short reads were normalized to 100x uniform coverage using BBnorm from the BBmap suite [20] and then subjected to guided de novo genome assembly using SPAdes (version 3.14.1) [21] with the *--careful* option. The guided SARS-CoV-2 genome assembly is achieved by using a reference genome with the –trusted-contigs option of SPAdes. The reference SARS-CoV-2 genome for the guided assembly was retrieved from UCSC [22]. The quality of the assemblies was assessed using QUAST (version 5.0.2) [23]. By using the guided approach, the vast majority of the assemblies was complete. For the few assemblies that were not complete, the MEDUSA scaffolder was deployed in order to complete the assemblies [24]. The short reads were also mapped to the SARS-CoV-2 genome retrieved from UCSC using BWA (version 0.7.17) [25] in order to further assess the quality of the virus sequencing and visually inspect coverage and potential virus mutations.

The complete de novo genome assembly and assessment procedure can be found online (https://github.com/moulos-lab/greek-covid19-assembly). The bioinformatics analysis was done using the computational infrastructure of the Center of New Biotechnologies & Precision Medicine (pMedGR), School of Medicine, National and Kapodistrian University of Athens, Greece (https://www.precisionmedicine.gr/).

### Sequence alignment and phylogenetic analysis

The dispersal patterns of SARS-CoV-2 in Greece were investigated by means of phylogenetic analysis. Classification of SARS-CoV-2 sequences in different lineages was implemented in the pangolin webtool (https://cov-lineages.org/pangolin.html). Our data consisted of: i) five datasets of 3,000 randomly selected sequences until September 30, 2020 sampled from the GISAID database plus the best hits identified by a BLAST search using as queries all sequences of our study population against the GISAID database sampled at the same time period, and ii) a dataset of 15,000 randomly selected sequences sampled until September 30, 2020 plus the best hits identified by a BLAST search. We collected 173,991 sequences from GISAID until 30^th^ of September, 2020 and created a Blast Database. We queried the 389 sequences and set a threshold to report the first 50 best hits (ranked by e-value and bitscore). BLAST reported pairwise 20,517 hits matching at different regions. These hits correspond to 2,059 unique sequences. The BLAST search was performed using only the coding region of SARS-CoV-2 (29,410 nucleotides). The total size of unique sequence datasets after the inclusion of the best hits, the random sets of 3,000 sequences and our study population, were 5,039 (dataset 1), 5,036 (dataset 2), 5,038 (dataset 3), 5,039 (dataset 4), and 5,036 (dataset 5). The size of the dataset including the 15,000 randomly selected sequences was 16,919 unique sequences. Analysis was performed without taking into account the classification of SARS-COV-2 into lineages.

Multiple sequence alignments were performed using the multithreaded version of MAFFT program [26]. This was run using XSEDE, available by the cyberinfrastructure for phylogenetic research (The CIPRES Science Gateway, version 3.3, https://www.phylo.org/), and the infrastructure at pMedGR.

Phylogenetic analyses were carried out by the maximum likelihood (ML) method using the IQ-Tree (version 2.1.1) [27] and the FastTree (version 2) programs [28]. Due to constrains in computation time, phylogenetic trees for sequence alignments larger than 3,000 sequences were imputed using FastTree. Phylogenetic analysis was performed using the best-fit nucleotide substitution with the ModelFinder and the Akaike Information Criterion (AIC) as implemented in IQ-Tree [27]. The nucleotide substitution model selected more often was the GTR+I+G4. For FastTree the GTR+G4 was selected as the nucleotide substitution model. All runs were performed at the CIPRES Science Gateway (version 3.3) and the infrastructure at pMedGR.

The resulting phylogenies were visualized and annotated by the FigTree (version1.4) and the Dendroscope (version 3.7.2) programs.

### Estimation of the number of imported SARS-CoV-2 infections

To infer the dispersal patterns of SARS-CoV-2 (i.e., the number of imported infections versus the within country infection events during different time periods), we performed phylogeographic analysis on all the 5 datasets of 5,036-5,039 sequences. Phylogeographic analysis was performed on the ML tree reconstructed by phylogenetic analysis conducted on the FastTree program in the previous step of analysis. Specifically, we estimated the number of SARS-CoV-2 infections (viral migration events) between different geographic areas/countries around the world and Greece (imported infections) during three time periods: between i) February 29 and March 31, 2020, ii) April 1 and June 30, 2020, and iii) July 1 and September 29, 2020. Additionally, we estimated infections occurring locally (local infections) between individuals for whom viral samples were taken at these three time periods. The viral migration events were quantified between the different geographic areas/countries by character reconstruction using the criterion of parsimony as implemented in PAUP*4.0 [29].

We assessed whether the inferred migration events (imported or local infections) were different from those expected by chance (panmixis). Hypothesis testing was performed by character reconstruction using the criterion of parsimony on the Mesquite (version 3.61) program [30].Further details on the methodology of viral migration event estimation have been described in detail elsewhere [31-34].

## Results

Our study data comprised of 389 unique full-genome SARS-CoV-2 sequences, of which 280 were newly generated and 109 were available on the GISAID database, collected in Attica until December 1, 2020 [35]. The vast majority of our samples had been collected in Attica (N=353, 90.7%). To investigate the patterns of SARS-CoV-2 infection in the areas of Northeast Greece and Thessaly, where virus surges were reported in March and May, respectively, we analysed 17 samples drawn from Alexandroupoli, Kavala, Komotini and Xanthi in Northeast Greece and 13 samples from the Nea Smirni area in Larissa, Thessaly. A few samples (N=4) analyzed as part of routine diagnostic testing in Attica were also available from 3 Aegean islands.

As mentioned in materials and methods section, sampling process comprised three time periods. The samples included in the study were as follows: 156 of 1,565 diagnosed cases (10%) for the first period, 101 of 1,873 cases (5.4%) for the second period, and 132 of 15,869 cases (0.8%) for the third period. The lower proportion for the third period was due to the number of tests performed increasing gradually with time (i.e., the average number of tests per month was approximately 10x higher in the third versus the first period), suggesting that the last period was not underrepresented in our sample.

The results of the classification of viral sequences into lineages, as estimated using the pangolin program, are shown in **Table 1**. The most frequent lineages were B.1.1 (European lineage; 40.6%), B.1.1.152 (Russian lineage; 19.5%), B1.1.38 (the UK lineage; 11.8%), B.1 (a European lineage that corresponds to the spring outbreak in Italy; 5.7%), B (basal lineage from China with many global exports; 4.4%) and B.40 (lineage dominant in the UK and Australia; 5.1%), while the A lineages originally detected at the early stages of the pandemic in Asia were present at low frequencies in Greece (A2: 0.5%; A5: 0.8%).

**Table 1.** Lineages of the study sequences per time period.

| **Time period** | **First** | **Second** | **Third** | **Total** | **Clustered<sup>1</sup>** |
| --- | --- | --- | --- | --- | --- |
| **Lineage** | Number of sequences (%) | Number of sequences (%) | Number of sequences (%) | Number of sequences (%) | Number of sequences (%) |
| A.2 | 2 (1.28) | - | - | 2 (0.51) | - |
| A.5 | 3 (1.92) | - | - | 3 (0.77) | - |
| B | 14 (8.97) | 3 (2.97) | - | 17 (4.37) | - |
| B.1 | 13 (8.33) | 2 (1.98) | 7 (5.3) | 22 (5.66) | - |
| B.1.1 | 81 (51.92) | 50 (49.5) | 27 (20.45) | 158 (40.62) | 81 (36.65) |
| B.1.1.1 | 1 (0.64) | - | 1 (0.76) | 2 (0.51) | - |
| B.1.1.38 | 5 (3.21) | 13 (12.87) | 28 (21.21) | 46 (11.83) | 44 (19.91) |
| B.1.1.70 | - | - | 1 (0.76) | 1 (0.26) | 1 (0.45) |
| B.1.1.100 | - | - | 1 (0.76) | 1 (0.26) | 1 (0.45) |
| B.1.1.102 | - | - | 1 (0.76) | 1 (0.26) | 1 (0.45) |
| B.1.1.145 | 1 (0.64) | - | - | 1 (0.26) | - |
| B.1.1.152 | 8 (5.13) | 23 (22.77) | 45 (34.09) | 76 (19.54) | 75 (33.94) |
| B.1.1.237 | - | 1 (0.99) | 2 (1.52) | 3 (0.77) | 3 (1.36) |
| B.1.1.291 | - | - | 9 (6.82) | 9 (2.31) | 9 (4.07) |
| B.1.1.315 | - | - | 1 (0.76) | 1 (0.26) | 1 (0.45) |
| B.1.5 | 4 (2.56) | 1 (0.99) | 2 (1.52) | 7 (1.80) | - |
| B.1.22 | - | 1 (0.99) | - | 1 (0.26) | - |
| B.1.36 | - | - | 3 (2.27) | 3 (0.77) | - |
| B.1.36.6 | - | - | 1 (0.76) | 1 (0.26) | - |
| B.1.98 | 2 (1.28) | - | - | 2 (0.51) | - |
| B.1.160 | - | - | 1 (0.76) | 1 (0.26) | - |
| B.1.177 | - | - | 1 (0.76) | 1 (0.26) | - |
| B.1.255 | 4 (2.56) | - | - | 4 (1.03) | - |
| B.1.319 | - | - | 1 (0.76) | 1 (0.26) | - |
| B.3 | 2 (1.28) | - | - | 2 (0.51) | - |
| B.4 | 1 (0.64) | - | - | 1 (0.26) | - |
| B.28 | 1 (0.64) | - | - | 1 (0.26) | - |
| B.39 | 1 (0.64) | - | - | 1 (0.26) | - |
| B.40 | 13 (8.33) | 7 (6.93) | - | 20 (5.14) | 5 (2.26) |
| **Total** | **156 (100)** | **101 (100)** | **132 (100)** | **389 (100)** | **221 (100)** |

<sup>1</sup> Distribution of sequences found within the largest local cluster per lineage.

To investigate the patterns of SARS-CoV-2 pandemic dispersal in Greece, we performed phylogenetic analyses on 5 different datasets, including as reference a random collection of globally sampled sequences and the best-hits identified by BLAST. The selection of sequences was performed over all SARS-CoV-2 lineages to avoid any biases stemming from lineages classification. Phylogenetic analyses on the different alignments revealed similar results, with at least 70 genetically distinct viruses identified as independent introductions in Greece. This number corresponds to the number of sequences (N=63) not falling within phylogenetic clusters with other sequences from Greece, hereafter named singletons, as previously reported [10], plus the number of clusters (N=7) comprising sequences from Greece. Given that our sample pool corresponds to 10% of the diagnosed cases and, also, that the actual number of SARS-CoV-2 infections should be severely underdiagnosed, the different lineages introduced to Greece should be higher than our estimation. The characteristics of SARS-CoV-2 phylogenetic clusters are shown in **Figure 1**, where, in addition to the 63 sequences that were not associated with onward transmission in Greece, we found several small clusters consisting of 2 to 6 sequences and two larger ones of 31 and 221 sequences (**Figure 2A-C**). The second largest cluster included 31 identical sequences sampled at the early stage of the pandemic from March 10 to 31, 2020 in Greece. The largest cluster consisted of 37 (16.7%; first period), 72 (32.6%; second period) and 112 (50.7%; third period) sequences collected during the respective sampling periods (**Figure 2C**). Notably, samples from the second and third period dominated in the largest local cluster. Furthermore, this cluster was underpinned by high levels of phylogenetic support (Shimodaira-Hasegawa, SH-support > 0.9) and was similarly detected in the phylogenetic tree performed using the random sampling of 15,000 GISAID sequences (SH-support > 0.85). The proportion of lineages for the sequences within the largest local cluster is shown in Table 1.

**Figure 1.**
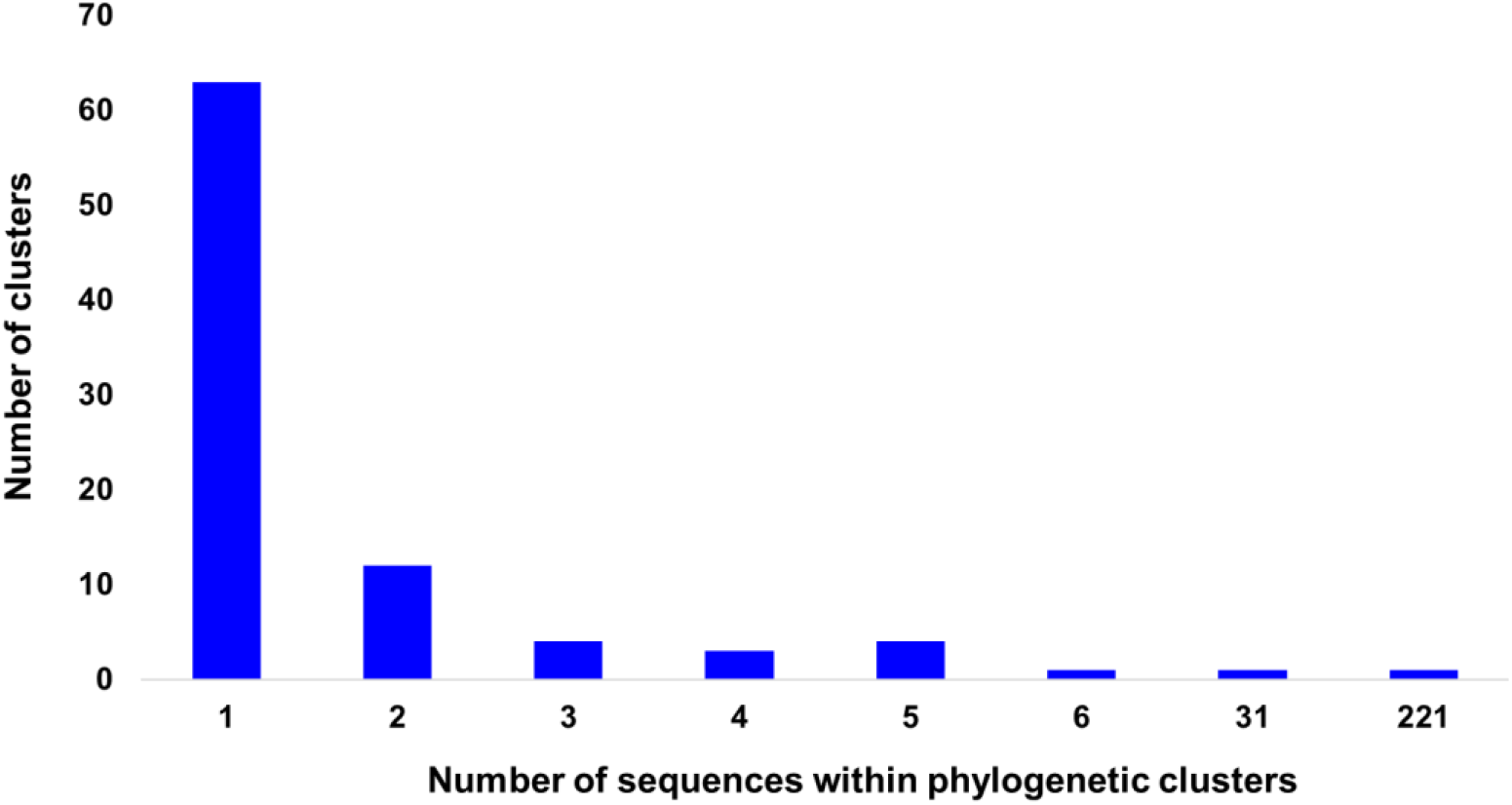
Distribution of number of sequences per phylogenetic cluster in Greece. The horizontal axis indicates the number of sequences within clusters and the vertical axis the number of the corresponding clusters.

**Figure 2.**
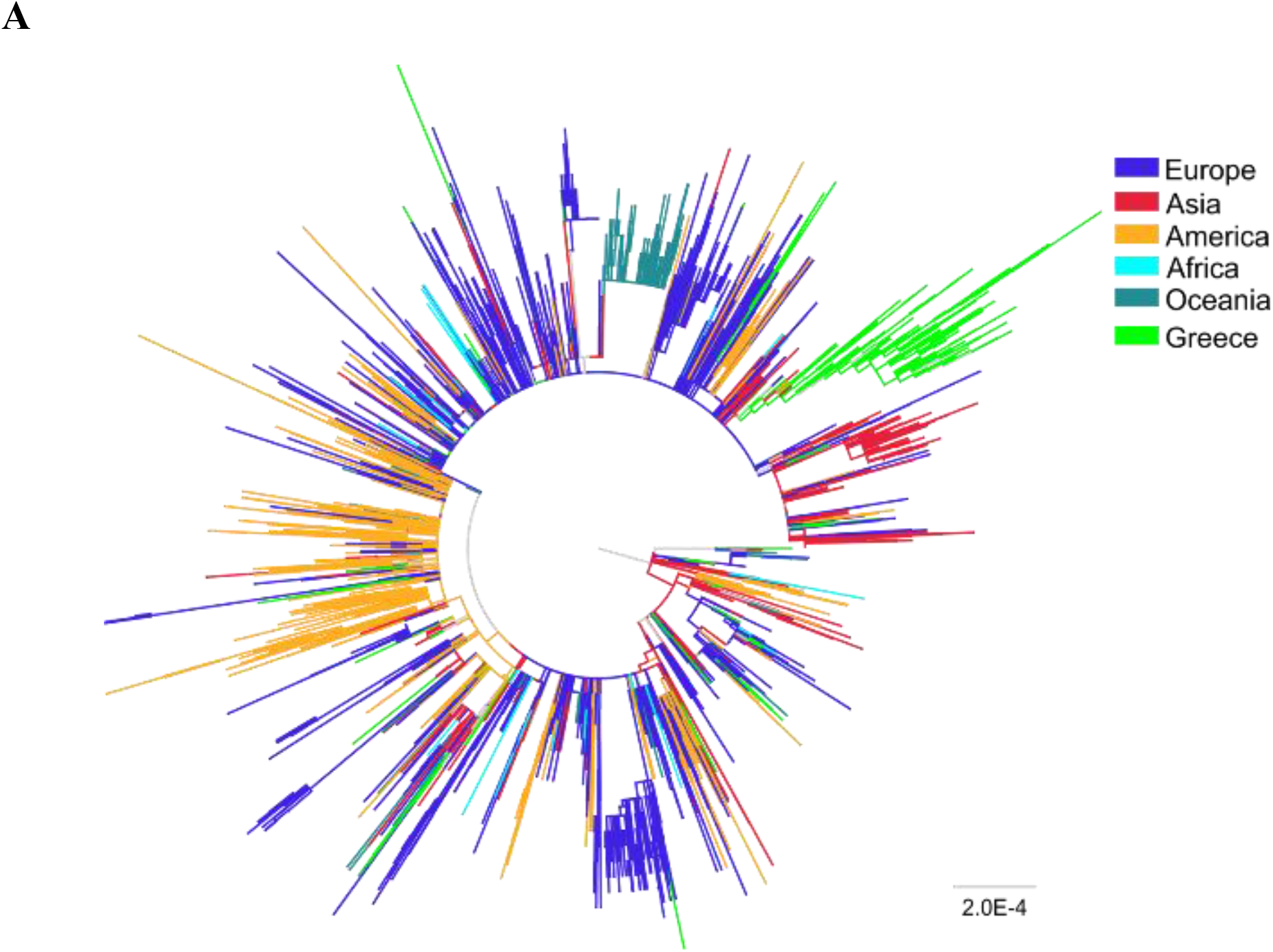
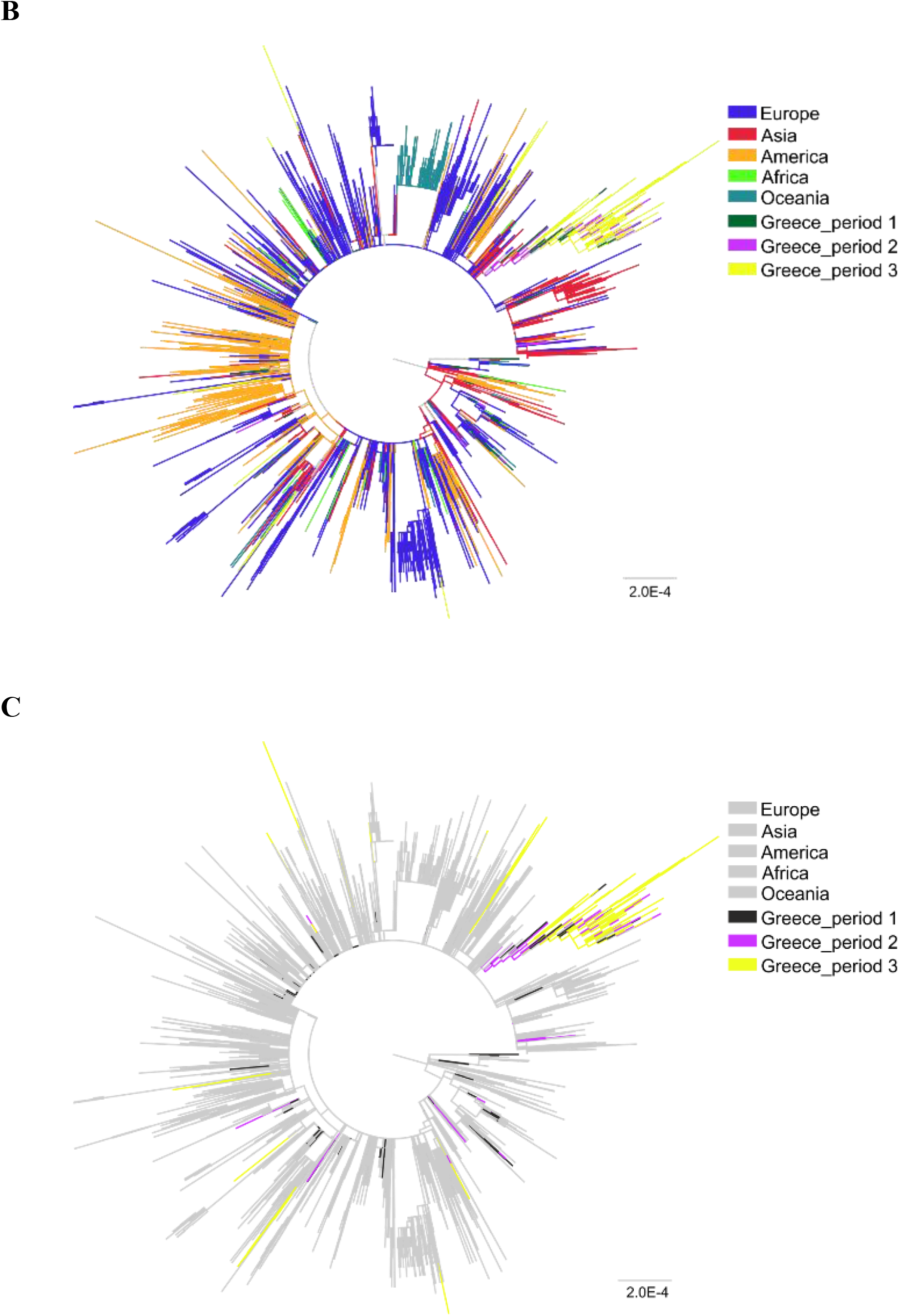
Unrooted phylogenetic tree estimated by FastTree (version 2) of SARS-CoV-2 sequences from Greece (N=389) and a global reference dataset N=4,647). **A**. All sequences from Greece are colored in light green. **B**. Sequences from Greece are marked in dark green (sampling period 1: 29/02/2020-31/03/2020: no travel restrictions) purple (sampling period 2: 01/04/2020-30/06/2020: travel restrictions) and yellow (sampling period 3: 01/07/2020-29/09/2020: lifting of travel restrictions). **C**. Sequences from Greece sampled from different time periods are shown in different colors and all reference sequences are shown in grey.

The putative numbers and sources of virus importation during the three time periods were inferred by means of phylogeographic analyses. The patterns of SARS-CoV-2 importation differed greatly between the three time periods: the proportion of imported infections peaked during the first period (mean value over the 5 datasets: 41%), while it remained low in the second (mean value over the 5 datasets: 11.5%) and third (mean value over the 5 datasets: 8.8%) periods (**Figure 3A**). The numbers of imported infections were similar across the different datasets and matched the proportion of imported cases reported from SARS-CoV-2 surveillance (**Figure 3A**). The corresponding figures were 31.2%, 15.5% and 13.8% for the three periods, respectively (**Figure 3A**). Implementation of travel restrictions and quarantine measures were applied in the middle of March, causing a decline in international arrivals, and were maintained until June (**Figure 3B**). Notably, the proportion of imported infections remained low after the lifting of restrictions on international travel implemented on July 1 in Greece (**Figure 3B**), and, although a virus surge was detected in August, it was not associated with an increased proportion of imported infections (**Figure 3C**).

**Figure 3.**
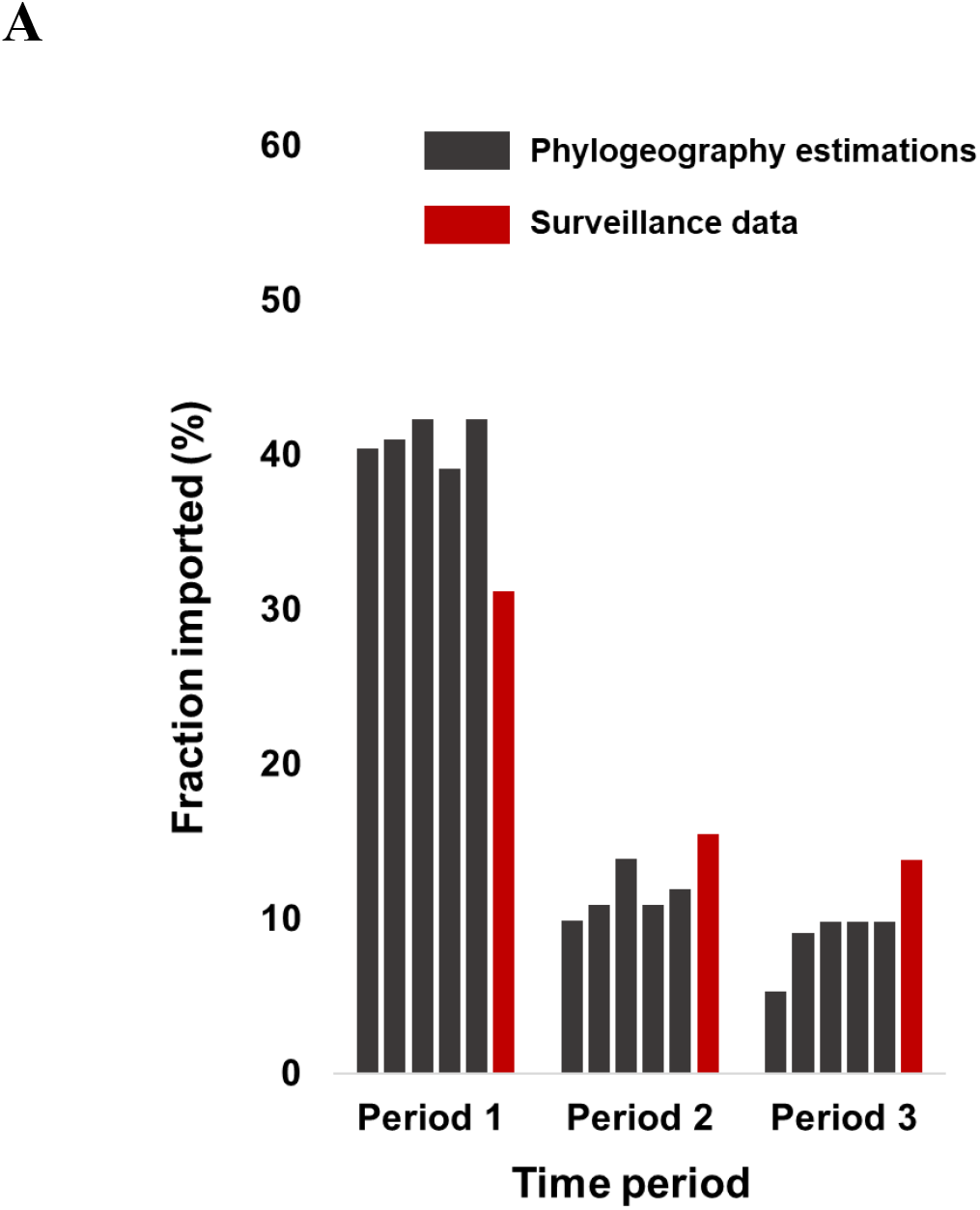
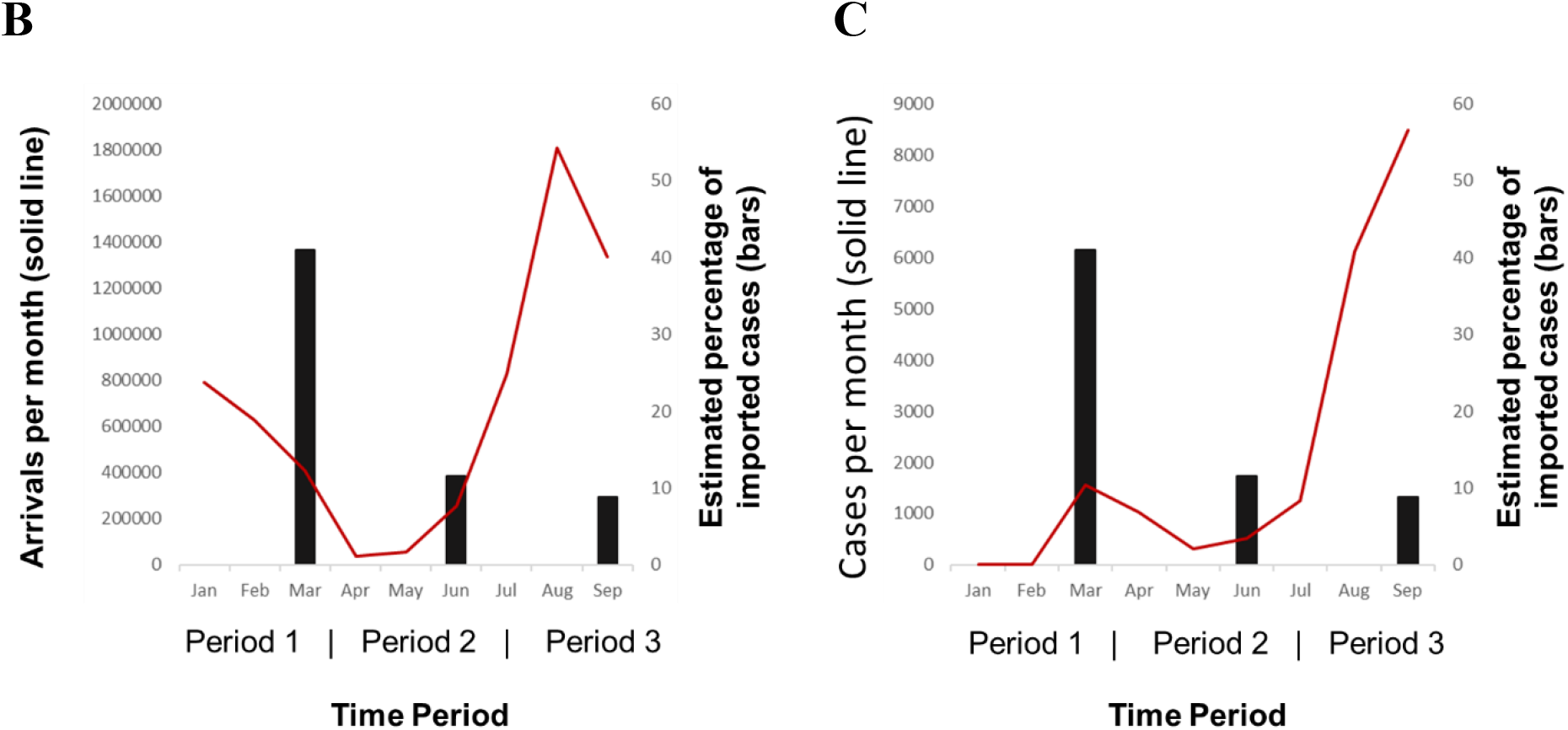
Proportion of virus importation estimated by phylogeographic analysis over the three sampling periods (sampling period 1: 29/02/2020-31/03/2020: no travel restrictions; sampling period 2: 01/04/2020-30/06/2020: travel restrictions; sampling period 3: 01/07/2020-29/09/2020: lifting of restrictions). **A**. Proportions of virus importation inferred by phylogeographic analysis using 5 different datasets (black) and surveillance data (red). Black bars indicate the proportion of virus importation inferred by phylogeographic analysis (mean value estimated from the 5 different datasets) in combination with **B**. the number of international arrivals per month (red line) and **C**. the number of SARS-CoV-2 cases per month in Greece (red line).

To investigate the significance of the pattern of virus importation, we compared the previous estimates with the expected proportions of imported infections under a scenario of panmixis. Our analysis suggested that the estimated proportions of imported infections were much lower than those expected by chance (**Supplemental Figure**), even during the first period (p<0.001); however, the differences were more pronounced in the second and third periods (p<0.001) (**Supplemental Figure**). These findings suggest that local transmission eventually dominated during the SARS-CoV-2 pandemic, but was less pronounced at the early stages, when travel restrictions had not yet been implemented.

The putative geographic origin of the imported infections showed that the majority originated from Europe and specifically from the UK (23 out of 43 imported cases; 53.5%) in the first period (**Table 2**). However, a considerable number of transmissions originated from non-European country (17 out of 43 imported cases; 39.5%) (**Table 2**). Subsequent analysis revealed that these cases were imported from America and Asia. During the second period, most of the imported infections were inferred to have originated from non-European country (8 out of 9; 88.9%) and the rest from the UK (1 out of 9; 11.1%) (**Table 2**). During the third period, half of the imported cases were traced to countries outside Europe and the remaining 33.3% and 16.7% were from the UK and Denmark, respectively (**Table 2**). According to the surveillance data, the highest number of imported SARS-CoV-2 cases were from the UK, at 36.1% and 16.8% for the first and second periods, respectively, proportions that were similar to those estimated by phylogeographic analysis. No information about the origin of potential imported cases was available for the third period since virus screening was performed at the entry sites and the putative origins of the imported cases that remained undiagnosed was unknown.

**Table 2.**
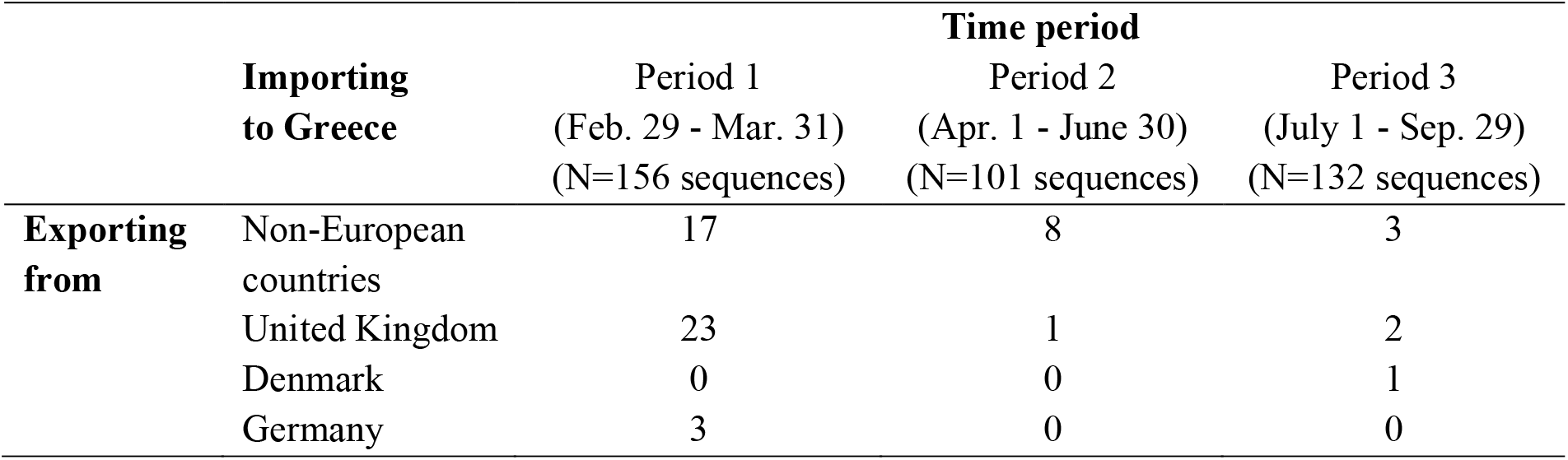
Estimated number of imported cases (migration events).

## Discussion

In the current study, using SARS-CoV-2 genomes from three distinct time periods, we showed that imported lineages were responsible for 41% of transmissions during the first pandemic wave in Greece. Moreover, we found that levels of virus importation significantly decreased during the period of travel restrictions and quarantine measures and, notably, remained low even after the opening of borders during the three months of peak tourist season. Our results were robust across different reference datasets and correlated strongly with the surveillance data regarding both the proportion of imported infections and the putative origin of SARS-CoV-2 lineages. Our findings suggest that imported infections dominated at the early stage of the pandemic before the implementation of travel bans.

More importantly, we found that virus importation remained low and did not substantially contribute to SARS-CoV-2 onward transmission even after the lifting of travel restrictions. Since July 1, 2020, all incoming travelers, including Greek citizens, need to have completed a passenger locator form (PLF) 48 hours before entering Greece. Health screening procedures have been put in place at airports and other ports of entry, where targeted testing has been performed guided by an artificial intelligence algorithm termed EVA. The algorithm combines information from previous tests performed at entry points in the country, as well as data obtained from the PLF creating an importation risk profile for each visitor according to country of travel origin. Health authorities can utilize this profile to determine border molecular testing prioritization, thus enhancing public health protection. Risk assessment for all countries was continuously performed daily and measures were modified accordingly, such as when entry restrictions were tightened for some countries when an increase in the number of SARS-CoV-2-positive cases was observed (a negative SARS-CoV-2 PCR test being required for entry). Additional public health measures were implemented (social distancing, local lockdowns, compulsory use of face masks in public spaces, etc.) locally or nationwide as necessary by local and national authorities.

Our study suggests that the impact of travelers to SARS-CoV-2 local transmission in Greece was low during the summer. To our knowledge, this is one of the few molecular epidemiology studies showing that the lifting of travel restrictions after the first pandemic wave was not associated with onward transmission driven by imported SARS-CoV-2 cases. This was most likely due to virus screening at entry points and public health measures implemented during the summertime and afterwards, which helped to control virus spread in the community. Notably, except for few islands (i.e., Paros, Mykonos), no virus surges were detected during the summer period in Greece and the effective reproductive number R remained around 1.1 −1.2 during this period (National Public Health Organization; unpublished data).

Early importation events observed during the first period resulted in large clusters only in one case; local clusters, that potentially have been ignited after the lifting of travel restrictions, may have remained undetected in our study. However, if this hypothesis were true, we would expect to have observed a high number of singleton (imported) infections, which was not the case. This suggests that the former hypothesis does not provide the most plausible explanation of the SARS-CoV-2 dispersal pattern during the third described period in Greece. Moreover, although our samples were not collected at tourist destinations, they were drawn from Attica, where almost 40% of the total Greek population resides, and which fuels tourism in these destinations during the summertime. Therefore, if new strains were associated with high levels of local transmission, we should have been able to detect them through our sampling.

Our findings are similar to those of previous studies in Europe and the Americas showing that the levels of imported infections declined after the implementation of travel restrictions during the first pandemic wave [10]. The scale of virus importation in Greece was in accordance with that in Boston, USA prior to March 28, 2020 (approximately 35%) and thereafter, when travel restrictions were implemented (median 9.3%) [7]. We also showed that the majority of strains during the first pandemic wave was imported from Europe, and specifically from the UK, but a significant proportion of virus importation originated in non-European countries. This pattern matched the origin of cases associated with travel during the first phase, suggesting that, although phylogeographic accuracy can be compromised due to potentially non-representative sequencing, in our analyses the putative origin of imported cases estimated by phylogeography matched that estimated by surveillance data.

Our study has several limitations. Our sampling was not representative and was not performed across Greece. On the other hand, as discussed above, our analysis was based on 389 full-length genomes collected at different time points from Attica, suggesting that our results reflect a large proportion of the population. Moreover, our study data included a dense sample of diagnosed cases during the first and second phases; sampling proportion was lower in the third period, but this was due to the enhanced testing performed over time. Regarding the putative limitation of non-sampling from tourist destinations during the third phase, if SARS-CoV-2 was continuously transmitted from viral lineages imported during the summertime in Greece, we would be able to detect them in Attica residents, a large proportion of whom visit many different places in Greece during the summertime. Our findings suggest that imported cases did not contribute substantially to SARS-CoV-2 local spread between July and September 2020. Importantly, our results on the effects of virus importation correlated with those estimated from surveillance data, thus enhancing the robustness of our findings. We should note that our findings are relevant to the summer period in the Mediterranean region and may not be generalizable for areas with different climatic conditions.

In conclusion, our molecular epidemiology study showed that the estimated proportion of imported cases during the first pandemic wave in Greece was 41% and that virus screening and public health measures after the lifting of travel restrictions prevented SARS-CoV-2 onward transmission from imported cases during summer 2020. These findings provide important insights on the efficacy of targeted public health measures and have important implications regarding the safety of international travel during a pandemic.

## Data Availability

The data will be made available in public databases.

## Acknowledgements

We acknowledge support of the next generation sequencing and bioinformatic analysis by the project “The Greek Research Infrastructure for Personalised Medicine (pMED-GR)” (MIS 5002802) which is implemented under the Action “Reinforcement of the Research and Innovation Infrastructure”, funded by the Operational Programme “Competitiveness, Entrepreneurship and Innovation” (NSRF 2014-2020) and co-financed by Greece and the European Union (European Regional Development Fund) and the Uni-Pharma S.A. We also acknowledge support of National Public Health Organization, the Greek Shipowners’ Social Welfare Company SYN-ENOSIS and the Public Benefit Foundation “John S. Latsis”.

## Supplemental material

**Supplemental figure.**
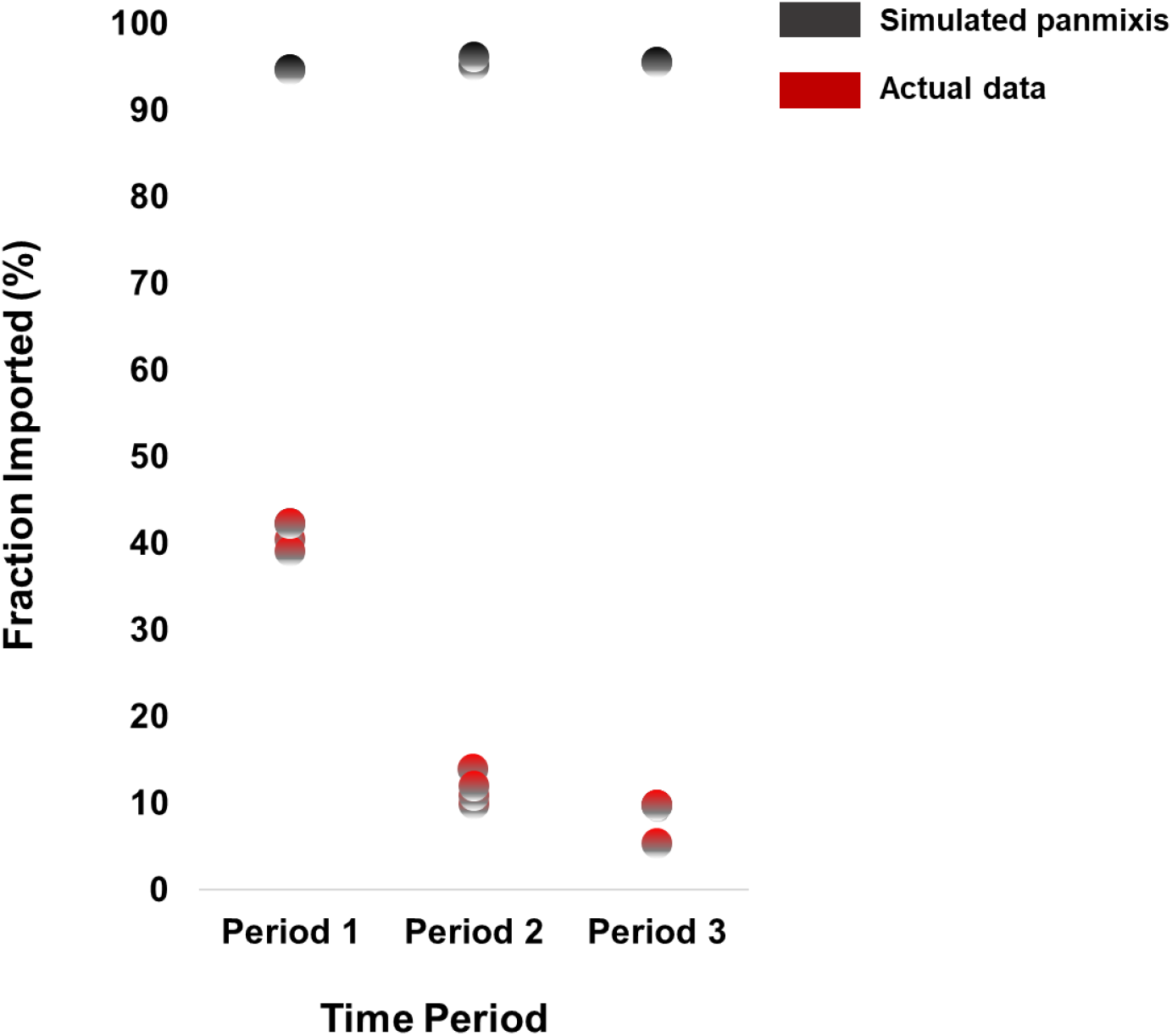
Proportion of virus importation estimated by phylogeographic analysis over the three sampling periods (sampling period 1: 29/02/2020-31/03/2020: no travel restrictions; sampling period 2: 01/04/2020-30/06/2020: travel restrictions; sampling period 3: 01/07/2020-29/09/2020: lifting of travel restrictions) (red dots) and after simulations of a scenario of panmixis (black dots).

